# Impact of COVID-19 Pandemic on Utilization of Facility-Based Essential Maternal and Child Health Services in North Shewa Zone, Ethiopia

**DOI:** 10.1101/2022.01.10.22268794

**Authors:** Chalachew Bekele, Delayehu Bekele, Bezawit M. Hunegnaw, Kimiko Van Wickle, Fanos Ashenafi, Michelle Korte, Christine Tedijanto, Lisanu Tadesse, Grace J. Chan

**Affiliations:** Birhan HDSS, St. Paul’s Hospital Millennium Medical College, Addis Ababa, Ethiopia; Department of Obstetrics and Gynecology, St. Paul’s Hospital Millennium Medical College, Addis Ababa, Ethiopia; Department of Pediatrics and Child Health, St. Paul’s Hospital Millennium Medical College, Addis Ababa, Ethiopia; Department of Epidemiology, Harvard T.H. Chan School of Public Health, Boston, MA, USA; Department of Global Health and Population, Harvard T.H. Chan School of Public Health, Boston, MA, USA; Francis I. Proctor Foundation, University of California San Francisco, San Francisco, CA, USA; HaSET MNCH research program, Addis Ababa, Ethiopia; Division of Medical Critical Care, Boston Children’s Hospital, Harvard Medical School Boston, MA, USA

## Abstract

**Introduction:** Ethiopia registered its first case of COVID-19 on March 13, 2020. We aimed to assess maternal, newborn, and child health care (MNCH) utilization during the first six months of the COVID-19 pandemic, as well as potential barriers and enablers of service utilization from health care providers and clients.

**Methods:** Mixed study design was conducted as part of the Birhan Health and Demographic Surveillance System in Ethiopia. The trend of service utilization during the first six months of COVID-19 was compared to corresponding time and data points of the preceding year.

**Result:** Service utilization of new family planning visits (43.2 to 28.5/month, p = 0.014) and sick under five child visits (225.0 to 139.8/month, P = 007) declined during the initial six months of the pandemic compared to the same period in the preceding year. Antenatal and postnatal care visits, facility delivery rates, and child routine immunization visits also decreased although this did not reach statistical significance. Interviews with health care providers and clients highlighted several barriers to service utilization during COVID-19, including fear of disease transmission, economic hardship, and transport service disruptions and restrictions. Enablers of service utilization included communities’ decreased fear of COVID-19, and awareness-raising activities.

**Conclusion:** Provision of essential MNCH services is crucial to ascertain favorable maternal and child health outcomes. In low- and middle-income country settings like Ethiopia, health systems might be fragile to withstand the caseloads and priority setting due to the pandemic. Our study presents early findings on the utilization of MNCH services that were maintained except sick child and new family planning visits. Government leaders, policy makers, and clinicians who wish to improve the resilience of their health system will need to continuously monitor service utilization and clients’ evolving concerns during the pandemic to prevent increases in maternal and child morbidity and mortality.

**What is already known?:** Facility-based essential MNCH services utilization decreased during the initial phase of the pandemic and similarly facility-based healthcare utilizations were reduced in the 2014-2015 Ebola outbreak in west Africa.

**What are the new findings?:** Facility based essential MNCH services such as antenatal care, postnatal care, family planning, facility deliveries, routine immunization and repeat family planning utilization were maintained in the initial six month of the pandemic unlike other similar studies elsewhere.

**What do the new findings imply?:** In light of a pandemic, essential MNCH services such as antenatal and postnatal care, family planning, facility deliveries, repeat family planning and routine immunizations can be sustained in a health system. More attention may be given to better understand the reduction of sick under five visits. Further research can be conducted on the utilization of essential MNCH services on maternal and child health outcomes. Our results emphasize the importance of health systems and clinicians to sustain the resilience of their health system. Among those the Ministry of Health (MoH) directive to avail MNCH services in all facilities during the pandemic and the maturity level of some programs (Even though new family planning utilizers are limited, they know the benefit and would want to continue the repeat family planning utilization, benefits of facility delivery, routine immunization, antenatal care and postnatal care).

**Strengths and limitations of this study:** *Strengths of the study:*

✓ We present primary data on service utilization during the early months of the pandemic in an area of Ethiopia, one of the agrarian regions, which is generalizable to 80% of the country’s population.
✓ The mixed methods approach integrated both quantitative service utilization coverage data with sociocultural, contextual, and exploratory qualitative to better understand our findings and reasons for changes in service utilization.
✓ The study highlights success stories in community-based care and government leadership for key services like routine immunization that may benefit other settings. Limitation of the study

✓ Our study focused on service utilization and may not have been powered to detect significant differences. Furthermore, we focused on coverage of service utilization as the primary outcome rather than mortality or morbidity.
✓ We do not have detailed data on service provision (e.g., which services were restricted and for how long, in what manner).
✓ There is the potential of recall bias were possible limitation since qualitative data was collected three months later than the initial six months of the pandemic (March to August 2020).

## INTRODUCTION

The World Health Organization (WHO) declared coronavirus disease-2019 (COVID-19) a global pandemic on March 11, 2020^1^ and the first case of COVID-19 in Ethiopia was registered on March 13, 2020.^2^ Multiple preventive measures focusing on social distancing and wearing masks were undertaken.^3^ Some health facilities were assigned as COVID-19 isolation and quarantine centers, and many suspended conducting elective surgeries and select outpatient services. This increasing burden of managing COVID-19 on health facilities and health care providers leaves the health system overstretched, challenging its ability to operate effectively. As shown during the 2014-2015 Ebola outbreak in west Africa, when health systems are overwhelmed by outbreaks, mortality from vaccine-preventable and other treatable diseases can increase dramatically.^4,5^

Well-organized and prepared health systems can continue to provide equitable access to essential services throughout an emergency^6^, but health systems in developing countries are often fragile when affected by emergencies such as pandemics. Accordingly, the WHO advises that countries should identify and prioritize essential services like routine vaccination, reproductive health services including care during pregnancy and childbirth, and care of young infants and older adults in their efforts to maintain continuity of service delivery and make strategic shifts to ensure that increasingly limited resources provide maximum benefit for the population.^7^ The disruption of services and diversion of resources away from essential sexual and reproductive health care due to the prioritization of the COVID-19 response are expected to increase risks of maternal and child morbidity and mortality.^8^

The Ministry of Health (MOH) of Ethiopia focused on prevention and control of the pandemic. Possible shifts of the health workforce and the health system towards the COVID-19 response may contribute towards low utilization of routine services. For example, “half sit” policies (29 March 2020) decreased maximum occupancy on public transit, thus cutting the number of seats in half and increasing the cost of transportation when traveling within the region while transportation between regions was paused completely. Additionally, school closures (March 2020), declaration of state of emergency (08 April 2020), awareness campaigns about the pandemic, and case reports, both suspected and confirmed cases, may have all contributed to growing fear of exposure to COVID-19, especially for patients visiting health facilities.

A modelling study of essential maternal and child health interventions across 118 low- and middle-income countries over a 6-month period estimated reduction of services by 9.8–18.5% and 39.3–51.9% in the least and most severe scenarios, respectively,^9^ due to the pandemic. Service reductions have already borne out in several contexts. In China, health service utilization declined significantly after the outbreak and all indicators rebounded beginning in March, but most had not recovered to their pre-COVID-19 levels by June 2020.^10^ In Bangladesh, Nigeria, and South Africa, between March and May 2020, the utilization of basic essential MNCH services such as antenatal care (ANC), family planning (FP), and immunization reduced due to lockdowns that triggered fear of contracting COVID-19, shifts of health system focused on managing the pandemic, and resource constraints.^11^ During the early phase of the COVID-19 outbreak (March-April 2020) in Rwanda, utilization of ANC, deliveries, postnatal care (PNC) and immunizations significantly declined.^12^ Similarly, a study in western Ethiopia showed a significant reduction in mean utilization of ANC, health facility birth, FP, and newborn immunization services between March–June 2019 and March–June 2020.^13^

In Nepal, a qualitative study found that maternity services, immunizations, and supply of essential medicine were the most affected health services during the lockdown. Interruptions were mainly due to the closure of health services at local health care facilities, limited affordability, involvement of private health sectors during the pandemic, fears of COVID-19 transmission among health care workers and within health centers, and disruption of transportation services. Participants expressed frustrations on poor testing, isolation and quarantine services related to COVID-19, and poor accountability from the government at all levels towards health services continuation and management during the COVID-19 pandemic.^14^

To understand these effects COVID-19 pandemic in Ethiopia, the HaSET Maternal and Child Health Research Program assessed trends in MNCH care utilization from March 2019 to August 2020 as well as health care providers’ and clients’ perceptions on the barriers to and enablers of service provision and utilization during the COVID-19 emergency. This study has paramount importance in filling the evidence gap on MNCH service utilization during COVID-19, both in the Ethiopian context and other low- and middle-income countries, to prevent significant damage to the gains achieved in such areas over the past several decades.

## METHODS

We conducted the study in eight health facilities, five health centers, and three hospitals (two public and one private), as part of the Birhan Health and Demographic Surveillance System (HDSS) in North Shewa Zone, Amhara Region, Ethiopia. Those facilities provide essential MNCH services for both rural (majority) and urban populations coming from HDSS catchment and non-catchment areas.

The health centers provide ANC, PNC, delivery, abortion, routine immunization (RI), integrated management of neonatal and childhood illness (IMNCI), and FP. Each health center also has a minimum of five service extension health posts mainly for FP and RI in each kebele (the lowest simplest administration unit), and each health post sends activity reports to health centers monthly. Two public hospitals (one primary and one referral) and the remaining one private general hospital also provide the above-mentioned essential MNCH services, except for RI, which is given mainly in health centers and catchment health posts.

Mixed qualitative and quantitative methods were employed. For the quantitative part of the study, a facility-based cross-sectional survey was conducted with MNCH healthcare providers to assess the impact of COVID-19 on essential MNCH service provision or utilization and provider-side barriers to service provision and utilization in Birhan catchment health facilities. Healthcare utilization time-series data from each facility was retrospectively collected and analyzed to understand the impact of COVID-19. In addition to this, a phenomenological qualitative design utilizing in-depth interviews was implemented to assess client and provider side barriers and enablers to service provision/utilization in Birhan catchment health facilities.

Birhan HDSS catchment health facilities’ medical records and monthly facility reports, interviews with health care providers working in the MNCH department, and interviews with women who delivered at home and facility, had ANC follow up, and who missed follow up were the data sources. All Birhan catchment health facilities were sampled for service statistics and health care providers who were working in essential MNCH departments and available at the time of visit were asked for respective sections. For the qualitative data, purposive sampling was implemented, and in-depth interviews were conducted until theoretical saturation was reached.

To assess health care providers’ perceptions on possible barriers to service utilization during the time of COVID-19, data were collected from interviews with health providers working in respective MNCH departments and facility and department heads. Retrospective facilities service statistics were collected over an 18-month period from March 2019 to August 2020 using Computer Assisted Field Editing (CAFE). Data was abstracted by uniformly structured questionnaires and entered to Open Data kit (ODK) collect and uploaded to ODK aggregate. The facilities’ monthly reports and medical registers data were collected separately. The monthly reports include services given in the health posts that are extension sites for the health center, but the facility registers are exclusively for services given in the health centers.

An interview guide with open-ended questions was used to elicit the qualitative information from informants. Face-to-face interviews were conducted in the facilities with women who visited facilities during COVID-19 and women who delivered at home. Women who missed an ANC follow up were interviewed by phone. With the permission of the respondent, all interviews were recorded, and all recorded data was transcribed for further analysis. To ensure the safety of the data collectors and participants, masks were worn, and social distancing practices were implemented during training and data collection from 2 – 20 November 2020.

The extracted data was exported to Stata 17.0 for analysis and the average MNCH services uptake was calculated each month to quantify the changes pre – COVID-19 (March to August 2019) and during the COVID-19 (March to August 2020) pandemic. For the purposes of analysis, March to August 2019 and March to August 2020 were considered as pre-COVID-19 and COVID-19 periods, respectively. To avoid the effect of missing and partially filled values, analogous months data from the same facility were excluded from the data analysis. Finally, an independent sample t-test was done to compare pre-COVID-19 and COVID-19 time months. This analysis was repeated for the initial two-months (March to April 2020) of the pandemic and the analogous period, March to April 2019, to examine changes in service utilization at the onset of the COVID-19 pandemic and a significance level of α=0.05 was used for all statistical tests.

In addition to the quantitative metrics listed above, English language transcript data was entered in Dedoose software for qualitative data analysis. After familiarization with the data, the content of the data was coded line by line for thematic analysis following a framework theory approach to describe and interpret health providers’ and communities’ perceptions on barriers and enablers to MNCH service provision. The framework approach involves using some pre-assigned themes to initially categorize data while also adjusting and iterating the coding scheme to accommodate newly emergent themes, sub-themes, and categories through inductive interpretation.^15^ Coded data was examined for potential relationships and themes were also assessed across relevant participant demographic categories to understand different user perspectives. Findings were described under pre-assigned and newly emerged themes.

### Ethic Statement

This study involves human participants and was approved by Ethics Review Board (IRB) of Saint Paul’s Hospital Millennium Medical college (SPHMMC) and Harvard T.H. Chan School of Public Health (HSPH) (IRB20-1574). To extract MNCH service statistics, permission was obtained from individual health facilities and individual verbal consent was obtained from respondents.

### Patient and Public Involvement

Meeting was restricted during protocol development and study period due to COVID-19 pandemic and it was not possible to involve clients or the public in the design, study, reporting and dissemination plans of our research.

## RESULTS

For the quantitative section of the study, data were abstracted from a total of eight health facilities (three hospitals and five health centers) and interviews with 103 healthcare providers working in the MNCH units of the facilities. In addition to these, ten facility or MNCH department heads and nine women (pregnant and delivered in the time of COVID-19) were asked open-ended questions.

Maternal health facility visits for ANC, PNC, facility delivery, and abortion-related services decreased in the time of COVID; however, we do not see a statistically significant change. The FP services utilization in the health centers and hospitals declined from 105.5 visits per month to 66.5 visits per month (p < 0.05) after the onset of the pandemic and within the subset of FP visits, repeat and unclassified FP visits significantly declined while new FP visits did not change. When combining health facilities with community health post data, the new FP services declined significantly from 43.2 visits per month to 28.5 visits per month (p = 0.029) but no significant changes in repeat, unclassified, and mean FP visits.

Declines in service utilization were found among sick child visits, which was defined as a facility visit for sick children under five years old. The mean number of IMNCI visits for sick children under 5 years old declined from 225.0 visits per month in 2019 to 139.8 visits per month in 2020 (p = 0.014). This significant relationship persists for two age stratifications of IMNCI visits (2 months to under 2 years, and 2 years to under 5 years). On the other hand, there was no significant change in child visits for routine immunizations, including BCG, OPV-0, pentavalent (DPT-HepB-HIP) and measles vaccinations.

“Ninety-one healthcare providers who were working in maternal, newborn and child health were asked about the client flow during COVID-19. Sixty-seven percent of the health care providers (HCPs) perceived that client flow decreased and 31% of them considered the same. Qualitative interviews also supported the observed decrease in client flow, with descriptions of sharper contractions in service utilization in the first couple of months after the onset of the pandemic but resumed to approximately normal levels over subsequent months. To explore the perception of lower service utilization during the initial couple of months of COVID-19 by clients of the health system and HCPs, data on the initial two pre-COVID (March to April 2019) months were compared with analogous COVID time months (March to April 2020) and there were no statistically significant changes in the number of visits for maternal and childhood visits overall, except sick child visit **(Supp Table 1)**.

### Barriers to service provision and utilization during COVID-19

Even though the essential MNCH service utilization was maintained; clients’ fear of acquiring the disease from the facility, travel restrictions, increased transportation cost due to the half seat order by the government, and fear of acquiring the disease on the way to the health facility were the main barriers for service utilization perceived by healthcare providers.

These secondhand perspectives from healthcare providers about the barriers that clients face was largely supported by qualitative interviews with clients. Fear of contracting the disease and lack of access to transportation are the most described barriers **(Supp table 2).** Particularly during the first few months after the onset of COVID-19 in Ethiopia and the imposition of travel restrictions and other public health measures like state of emergency, communities fear of acquiring the disease and high levels of public panic were barriers for facility-based services utilization. This fear extended to visiting facilities during the pandemic; community members were afraid of contracting the disease in crowded spaces, public transportation routes to facilities, or at facilities themselves from health care workers or other patients, particularly when they heard of COVID-19 cases present at facilities and this fear often resulted in delayed care-seeking. A woman said that *“I have postponed my follow up at that time for fear of acquiring the disease from health professionals and health centers. The same is true for other clients in our area, and some mothers have received their visit in private clinics as we perceived almost all staff were infected”*.

The economic hardship during COVID-19 prevented some clients from being able to pay for transportation due to transportation restriction with half seats and doubled transportation fee, and other direct or indirect costs of attending facilities. Clients described lacking money to purchase PPE and one HCP noted that the closed market movement affected people’s incomes, as reflected by patients delaying treatment until conditions are more severe or defaulting on treatment.

Lastly, bottlenecks on the health system side provided another barrier to service utilization. Facilities restricted some services at the beginning of the pandemic’s onset and clients were unable to access certain services or assumed that services were restricted even after they had resumed, as one HCP suggested. Additionally, multiple clients described fearing that they might be forcibly quarantined or presumed COVID-19-positive if they were to visit facilities and this fear deterred facility visits. HCPs described many challenges related to the under preparedness of the health system for managing suspected cases of COVID-19. Often these challenges manifested through physical infrastructure constraints and a shortage of guidelines for managing quarantine and isolation centers for suspected COVID-19 cases.

### Enablers of service provision and utilization during COVID-19

In terms of knowledge of COVID-19, all women had heard about the disease, but a few were in doubt about COVID-19 existence in the area which may be an enabling factor for facility-based service utilization. A client respondent said, “*I do not believe it exists, especially in our area. It might be real / exist in other areas/countries. They just suspect and take everyone into an isolation/quarantine center, but they are healthy and free of any signs and symptoms…*”. Some described that COVID-19 could not affect them because God and/or Mary will protect them, citing the importance of prayer as a protective measure.

Facility adaptations like training for healthcare providers, hand washing facilities, physical distancing, and awareness creation and health education which was given by local authorities increased the client awareness on COVID-19 prevention and facility-based service utilization through time **(Supp table 3).**

### Understanding Service Utilization Trends during COVID-19

The barriers and enablers highlighted in the interviews interact with each other dynamically, as depicted below. Certain barriers were more substantial than others, particularly fear of the disease, transportation access, and economic-related barriers, while certain pull factors encouraged facility visits, particularly over time as fear subsided, community awareness measures were undertaken, and facilities implemented adaptations to manage both COVID-19 and routine services **(Diagram 01).**

**Diagram 1:**
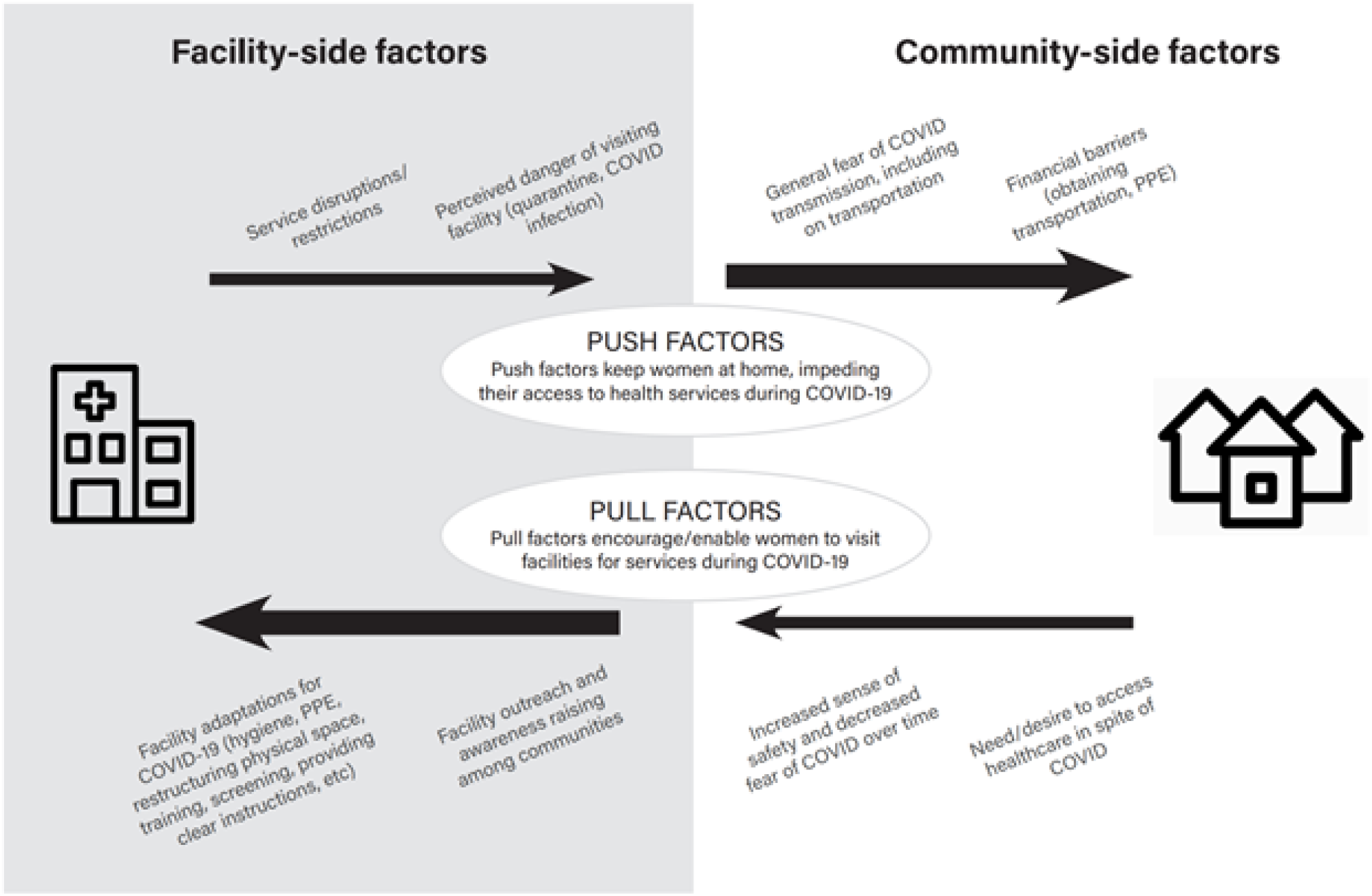
Enabling (pull) factors and barriers (push factors) for service utilization highlighted in the qualitative interviews.

## DISCUSSION

We examined the impact of the COVID-19 pandemic on essential MNCH service utilization by analyzing data from health facility records and healthcare providers and patients’ perspectives. In the context of already poor health outcomes, significant reductions in service utilization for maternal and child health may have substantial adverse impacts. A modelling study of 118 low- and middle-income countries estimated an additional 12,200-56,700 maternal and 253,500-1,157,000 child deaths using several hypothetical scenarios in which the coverage of essential maternal and child health interventions were reduced by 9.8-51.9% due to the pandemic over 6 months.^9^

For maternal health, FP, ANC, PNC, facility delivery, and abortion services utilization decreased, but the change was not significant during the initial six months of the pandemic. Globally, contraception services were shut down or not accessible,^16^ which was also observed in our study resulting in reduced family planning service utilization and a drastic drop in new family planning services. Service utilization for family planning was stable at health posts (community-based clinics that are an extension of health centers) suggesting that utilization of family planning services was more likely to occur when clinics are nearby without extensive travel.

For child health, the number of Integrated Management of Neonatal and Childhood Illness (IMNCI) visits, also referred to as sick child visits, significantly declined by 38%. It is possible that the decrease in child sick visits was related to COVID-19 prevention and control activities. The leading causes of under five years old children morbidity in Ethiopia,^17^ acute respiratory illness, fever and diarrhea may have decreased due to school closures (older siblings less exposed), limited interactions with peers in the community, spending more time indoors, mask-wearing at community gatherings, hand washing, physical distancing, and other personal protective equipment and practices. We found that RI remained stable during the initial six months of the pandemic, which was different than the findings from studies in Colombia, India, and Brazil where RI declined during the pandemic.^18-20^ The Ethiopian MOH prioritized RI, especially measles, during COVID-19. Ethiopia deploys health extension workers stationed at health posts for community-based services hygiene and sanitation, FP and RI which may have sustained accessibility to these services during COVID-19. It is worth nothing that the source of data for FP and RI in this study included aggregated data from the health management information system (HMIS) at health centers which may be less reliable than directly collecting data from hospital and health centers’ records other services like IMNCI and sick child visits. ^21^ In Ethiopia, during COVID, there was a marked reduction in supply chain distribution of vaccines implying that the RI coverage like decreased nationally.^22^

Overall, with the exception of RI, our findings are similar to other global studies that have also found declines in service utilization during the COVID-19 pandemic. A systematic review of eighty-one studies in twenty countries reported a reduction in health care utilization with a median 37% reduction in overall services, including 42% reduction for visits, 28% for admissions, 31% for diagnostics, and 30% for therapeutics between pandemic and pre-pandemic periods in the initial two months of the pandemic.^23^ There was also a reduction in utilization of basic essential MNCH services such as ANC, FP and RI in Bangladesh, Nigeria, Rwanda and South Africa between March and May 2020.^11,12^ In a semi-pastoralist area in western Ethiopia, there was a significant reduction in mean utilization of ANC, health facility births, FP, and RI.^13^

The reduction in service utilization was observed in the setting of the communities’ experiences of and perceptions toward COVID-19, including misinformation, misconceptions, and doubt. As the pandemic has progressed, service utilization may further decline as fear settles in. At the time of data collection, early in the pandemic, respondents largely described not feeling many tangible impacts of COVID-19 on their daily lives or consequences of lack of adherence to preventive measures, so they went about life as usual. This easing fear of COVID-19 may have enabled women to feel that they could safely attend services, but it also has important implications as the pandemic continues, particularly as cases in Ethiopia have risen substantially. Awareness and education campaigns are needed to produce actual behavior change. Moreover, communities’ belief that God may protect them from infection indicates the important role of engaging religious leaders as champions in behavior change campaigns. An additional key recommendation is systematically addressing misinformation and doubt to increase population compliance with preventive measures, particularly as Ethiopia faces a rising caseload, increasing prevalence of variants, and a stalled vaccine rollout that may take months or years to reach substantial population coverage. Less than 2% of the population has received at least one dose of COVID-19 vaccine.^24^

Barriers to maternal facility visits included women not wanting to bother anyone, lack of support from healthcare workers, influence of the media,^25^ lockdown, fear of contracting the disease,^26^ shift of focus towards pandemic, and resource constraints.^11^ In addition, women experienced fears of contracting the disease, economic hardship, and lack of access to transportation. Particularly during the first few months after the onset of COVID-19 in Ethiopia, there was an imposition of travel restrictions and other public health measures like state of emergency, and high levels of public panic. In addition, facilities restricted some services at the beginning of the pandemic and clients were unable to access certain services. Multiple clients described fearing that they might be forcibly quarantined or presumed COVID-19-positive if they were to visit facilities; this fear deterred facility visits. While we found that sick child visits and new family planning services were most affected by the pandemic, the declines among other essential services were not as significant indicating hope in service resilience and the ability to introduce rapid and substantial facility adaptations to maintain the health system (e.g., personal protective equipment, infection prevention measures, improved sanitation/hygiene).

### Strengths of the study

We present primary data on service utilization during the early months of the pandemic in an area of Ethiopia, one of the agrarian regions, which is generalizable to 80% of the country’s population. We leveraged an existing research network, HaSET MNCH research program (www.hasetmch.org), and established field site^27^ to rapidly collect data from all available sources. The mixed methods approach integrated both quantitative service utilization coverage data with sociocultural, contextual, and exploratory qualitative to better understand our findings and reasons for changes in service utilization. The study highlights success stories in community-based care and government leadership for key services like routine immunization that may benefit other settings.

### Limitation of the study

Our study focused on service utilization and may not have been powered to detect significant differences. Furthermore, we focused on coverage of service utilization as the primary outcome rather than mortality or morbidity. We do not have detailed data on service provision (e.g., which services were restricted and for how long, in what manner). There is the potential of recall bias were possible limitation since qualitative data was collected three months later than the initial six months of the pandemic (March to August 2020).

## CONCLUSION

Utilization of essential MNCH services is crucial to achieve favorable health outcomes. In the setting of developing countries like Ethiopia, health systems are often too fragile to withstand the direct increase in volume of patients and indirect health consequences of a pandemic. Our study presents early findings on the decline in the utilization of MNCH services especially in new family planning services and sick child visits. Further study is needed to assess the effect of service utilization decline on MNCH morbidity and mortality. To prevent worsening maternal and child morbidity and mortality as a result of the pandemic, resources are required by government leaders, policy makers, and clinicians to improve the resilience of their health system to continuously monitor service utilization, while at the same time engaging with providers and clients to understand and address their evolving concerns about MNCH service uptake.

## Data Availability

The datasets used and/or analyzed during the current study are available from the corresponding author on reasonable request.

## ABBREVIATIONS

CAFÉ: Computer Assisted Field Editing,
COVID-19: Coronavirus Disease - 2019,
DPT: Diphtheria Pertussis and Tetanus,
EOC: Emergency Operation center,
EPHI: Ethiopian Public Health Institute,
FP: Family Planning,
HC: Health Center,
HCP: Healthcare provider,
HDSS: Health and Demographic Surveillance System,
HepB: Hepatitis B,
HIP: Haemophilus influenzae,
HIV/AIDS: Human Immunodeficiency Virus/ Acquired Immune Deficiency Syndrome,
HP: Health post,
HSPH: Harvard School of Public Health,
IRB: Institutional Review Board,
MCH: Maternal and Child Health,
MNCH: Maternal, Newborn and Child Health,
MOH: Ministry of Health,
ODK: Open Data Kit,
PNC: Postnatal care,
RI: Routine immunization,
RMNCH: Reproductive, Maternal, Newborn and Children Health,
SPHMMC: Saint Paul Hospital Millennium Medical College,
W: Women,
WHO: World Health organization.

## ACKNOWLEDGEMENTS

We acknowledge the kind help and encouragement we received from the zone, woreda and study facilities. Our deepest gratitude goes to the women and health care providers who participated in this study and to all data collectors and supervisors for their dedicated work.

## GRANT/AWARD NUMBER

The research is co-funded through INV-006752 HaSET and INV-003612 ANC/PNC Innovations Platform HaSET Ethiopia Partnership.

---

**Table 1.** Comparing essential MNCH service utilization over six months between COVID-19 (Mar-Aug 2020) and analogous pre-COVID-19 (Mar-Aug 2019) periods.

| Visit Type | Mean number of visits/month over six months |  | t-statistic | p-value | Lower p-value<sup>2</sup> | Upper p-value<sup>2</sup> | Paired observations |
| --- | --- | --- | --- | --- | --- | --- | --- |
|  | 2019 | 2020 |  |  |  |  |  |
| **I. Maternal visit** | **376.3** | **321.2** | **1.11** | **0.270** | **0.865** | **0.135** | **48** |
| 1. Antenatal care | 208.9 | 181.7 | 0.79 | 0.433 | 0.784 | 0.216 | 40 |
| 2. Postnatal care | 26.6 | 19.8 | 1.44 | 0.155 | 0.922 | 0.078<br>* | 30 |
| 3. Facility delivery | 90.7 | 84.2 | 0.29 | 0.776 | 0.612 | 0.388 | 41 |
| 4. Abortion-related services | 11.8 | 9.8 | 0.56 | 0.578 | 0.711 | 0.289 | 34 |
| 5. Combined FP services in HP, HC and hospitals | 313.3 | 273.4 | 0.82 | 0.415 | 0.792 | 0.207 | 47 |
| 5.1 New FP services | 43.2 | 28.5 | 1.22 | 0.029 | 0.986 | 0.014<br>** | 47 |
| 5.2 Repeat FP services | 270.2 | 244.9 | 0.57 | 0.567 | 0.716 | 0.284 | 47 |
| 6. FP services in HCS and hospital | 105.5 | 66.5 | 1.99 | 0.051 | 0.974 | 0.026<br>** | 33 |
| 6.1 New FP services | 8.9 | 7.1 | 0.84 | 0.406 | 0.797 | 0.203 | 33 |
| 6.2 Repeat FP services | 96.5 | 59.3 | 1.03 | 0.046 | 0.977 | 0.023<br>** | 33 |
| 6.3 Unclassified FP services | 17.7 | 1.3 | 1.12 | 0.039 | 0.981 | 0.019<br>** | 26 |
| **II. Sick child visit (0-5years))** | **225.0** | **139.8** | **1.51** | **0.014** | **0.993** | **0.007**<br>*** | **46** |
| 1. MNCI Visit (< 2 months) | 10.8 | 7.7 | 0.82 | 0.412 | 0.794 | 0.206 | 46 |
| 2. IMNCI Visit (2 months – 2 year) | 101.6 | 50.4 | 1.68 | 0.009 | 0.996 | 0.004<br>*** | 46 |
| 3. IMNCI Visit (2 year – 5 year) | 111.6 | 81.8 | 1.15 | 0.034 | 0.983 | 0.017<br>** | 46 |
| **III. Routine Immunization visit** | **37.0** | **36.8** | **0.02** | **0.982** | **0.509** | **0.491** | **23** |
| 1. BCG Vaccine | 31.4 | 36.5 | -0.39 | 0.701 | 0.350 | 0.650 | 30 |
| 2. Oral Polio (0) Vaccine | 3.2 | 1.0 | 0.88 | 0.384 | 0.808 | 0.192 | 23 |
| 3. Pentavalent (DPT-HepB-HIP)<br>(all types) | 100.4 | 101.5 | -0.05 | 0.958 | 0.479 | 0.521 | 30 |
| 4. Measles – 1 | 10.3 | 27.5 | -1.99 | 0.051 | 0.026 | 0.974 | 30 |
| 5. Vitamin A Dose | 8.6 | 5.7 | 0.67 | 0.506 | 0.747 | 0.253 | 14 |
| **Other types of visits** |  |  |  |  |  |  |  |
| All visits | 2568.9 | 2606.7 | -0.05 | 0.956 | 0.478 | 0.522 | 48 |
| Adult Outpatient Visit | 2121.2 | 2239.7 | -0.17 | 0.868 | 0.434 | 0.566 | 44 |
| * p < 0.10 ** p < 0.05 *** p < 0.01, □ Lower-tailed test: mean number of visits H<sub>1</sub>: $\mu_{2019} < \mu_{2020}$, □□ Upper-tailed test: mean number of visits H<sub>1</sub>: $\mu_{2019} > \mu_{2020}$ | | | | | | | |

**Table 2.** Possible barriers to service utilization in the time of COVID-19 based on healthcare providers’ perception.

| Possible barriers to service utilization in the time of COVID-19 | Count | % |
| --- | --- | --- |
| 1. Fear of acquiring the diseases from the facility | 97 | 94 |
| 2. Travel restrictions | 90 | 87 |
| 3. Increased transportation cost (due to half sit order by the government) | 89 | 86 |
| 4. Fear of acquiring the disease on the way to the health facility | 86 | 83 |
| 5. Lack of transport to the HP/HC site | 72 | 70 |
| 6. Lack of PPE for clients | 67 | 65 |
| 7. Clients' perception of limited implementation of protective measures by<br>8. healthcare providers | 58 | 56 |
| 9. Healthcare providers advice to stay at home | 54 | 52 |
| 10. Limited-service hours or absence of health care workers | 17 | 17 |
| 11. Unavailability of ambulance | 7 | 7 |
| 12. Unavailability of healthcare providers in facilities to provide outreach<br>services. | 7 | 7 |
| Total eligible respondents | 103 |  |

