## Supplemental Tables for "Impact of COVID-19 Pandemic on Utilization of Facility-Based Essential Maternal and Child Health Services in North Shewa Zone, Ethiopia"

### **Supplementary Tables**

**Supplementary Table 1.** Comparing essential MNCH service utilization over two months between COVID (April- May 2020) and analogous pre-COVID (April-May 2019) periods.

| Visit Type | Mean number of visits/ months over six months | | t-statistic | p-value | Lower p-value | Upper p-value | Number of paired observations |
| --- | --- | --- | --- | --- | --- | --- | --- |
|  | 2019 | 2020 |  |  |  |  |  |
| I. Maternal visit | 331.3 | 327.3 | 0.05 | 0.959 | 0.520 | 0.480 | 16 |
| 1. Antenatal care | 198.0 | 176.5 | 0.39 | 0.702 | 0.649 | 0.351 | 13 |
| 2. Postnatal care | 17.8 | 20.7 | -0.30 | 0.768 | 0.384 | 0.616 | 9 |
| 3. Facility delivery | 85.6 | 87.0 | -0.04 | 0.970 | 0.485 | 0.515 | 14 |
| 4. FP related services | 84.1 | 95.1 | -0.38 | 0.712 | 0.356 | 0.644 | 10 |
| 5. FP related services (hospitals, HCs and HPs combined) | 289.4 | 227.8 | 0.86 | 0.398 | 0.801 | 0.199 | 16 |
| 6. Abortion-related services | 10.5 | 10.7 | -0.05 | 0.964 | 0.482 | 0.518 | 11 |
| II. Sick child visit *(0-5years*) | 201.0 | 126.6 | 1.68 | 0.103 | 0.948 | 0.052* | 15 |
| 1. IMCI Visit (< 2 months) | 7.3 | 3.3 | 1.21 | 0.235 | 0.882 | 0.118 | 15 |
| 2. IMNCI Visit (2 months – 2 year) | 105.0 | 77.4 | 1.54 | 0.134 | 0.933 | 0.067* | 15 |
| 3. IMNCI Visit (2 year – 5 year) | 89.7 | 45.9 | 1.43 | 0.164 | 0.918 | 0.082* | 15 |
| III. Routine Immunization visit | 41.0 | 38.1 | 0.16 | 0.875 | 0.563 | 0.437 | 7 |
| 1. BCG Vaccine | 28.0 | 36.9 | -0.40 | 0.695 | 0.347 | 0.653 | 10 |
| 2. Oral Polio (0) Vaccine | 3.4 | 1.0 | 0.61 | 0.556 | 0.722 | 0.278 | 7 |
| 3. Pentavalent (DPT-HepB-HIP) (all types) | 100.4 | 101.4 | -0.03 | 0.978 | 0.489 | 0.511 | 10 |
| 4. Measles – 1 | 5.6 | 28.3 | -1.45 | 0.163 | 0.082* | 0.918 | 10 |
| 5. Vitamin A Dose (any dose) | 6.0 | 1.0 | 0.87 | 0.419 | 0.791 | 0.209 | 4 |
| Other types of visits | | | | | | | |
| 4. All visits | 2031.9 | 2323.5 | -0.27 | 0.787 | 0.394 | 0.606 | 16 |
| 5. Adult Outpatient Visit | 1811.6 | 2147.5 | -0.29 | 0.773 | 0.386 | 0.614 | 14 |
| * p < 0.10 ** p < 0.05 *** p < 0.01, ^♱^ Lower-tailed test: mean number of visits H_1_: μ_2019_ < μ_2020,_ ^♱♱^ Upper-tailed test: mean number of visits H_1_: μ_2019_ > μ_2020_ | | | | | | | |

***Supplementary Table 2.*** *Themes and illustrative quotes on factors enabling community facility visits during COVID-19.*

| Themes | Illustrative Quotes |
| --- | --- |
| COVID-19 perception | |
| People are in doubt about COVID-19 existence in the area. | - I do not believe it exists, especially in our area. It might be real / exist in other areas/countries. They just suspect and take everyone into an isolation/quarantine center, but they are healthy and free of any signs and symptoms…. (W) - ….. I have never seen anyone with such a real problem in our area. We have heard about it on radio and TV, so I found it difficult to believe and I do not believe it is real (W). - There are huge gaps, misconceptions, and challenges in practical preventive practices. They even perceived that the disease may not be real. Clients recovered from COVID-19 without any sign and symptom disseminated the information to the community and based on that the community misconceived that the virus might not be real from the beginning (HCP). - Right now, the entire community members have no fear or concern about acquiring the disease ……. we are not concerned about client decrement related to COVID-19. Specially after the 5 months state of emergency was lifted things are returned to pre-COVID time, …. (HCP). |
| No/Low COVID-19 impact perception on daily life | - COVID was for outsiders not for us, it was for political issues, the machine for COVID test was false (W). - Has COVID-19 been affecting your life in any way? P: No nothing (W) - I do not think we are at risk because we are not getting out of home most of the time and living in rural areas without any contact (W). |
| Knowledge on transmission methods | - Crowding at one place like the market and public transportation(W). - She laughed. “Media expresses it well; we know well it is also an infected person who can transmit it …” it was not on her tip of tongue she encouraged simply to remember and told me freely “…contact, breathing” (W). - It can be transmitted through air/ breathing, shaking hands, kissing, contact with others and when face masks are not applied properly (W). |
| Facility adaptation | |
| Training provided to HCP | - …. there was continuous and repeated awareness creation on the preventive measures, how they apply it to prevent COVID-19… (HCP) - After the first case of COVID-19 was confirmed in our country, all health care providers including supportive staff were oriented about covid-19 and how to protect themselves and their clients (HCP). - Training was given for all health professionals by trained woreda health professionals, how the health professional can use mask and keeping distance, source of the virus’s transition and the like (HCP). |
| PPE use and social distancing | - All health workers have applied face masks and sanitizer while providing services (W). - Health professionals kept all PPE materials in place while serving clients (HCP). - Health professionals have put on their face masks, enforce clients to wear face masks during facility visits and hand washing soap has also been kept in place for clients (W). - We had arranged client sitting chairs at all departments to keep their social distance; we had assigned one personnel to educate and to keep their social distance (HCP). - We were giving care for patients face to face in and in close contact so far, but now we are providing two meters distance (HCP). |

***Supplementary Table 3.*** *Themes and illustrative quotes on perception of client flow and barriers of community facility visit.*

| Themes | Illustrative Quotes |
| --- | --- |
| Perception of client flow | |
| Client flow decreased initially and increased through time | - During my ANC visit, I have seen some clients receiving health services. At the beginning of coronavirus some people did not want to receive the services for fear of contracting the disease. So, client flow at that time has decreased (W). - Following of covid-19 positive case detection in the country, somewhat patient flow was decreased HCP). - During COVID-19 time, the patient flow has dramatically decreased at the beginning (HCP). - Right now, the entire community members have no fear or concern about acquiring the disease (HCP). - Becomes the same as pre-COVID-19 time since the state of emergency lifted (HCP). |
| Barriers for service utilization | |
| Fair of acquiring the disease in the facility | - You can have this risk at transport and at health facilities during service provision and from other clients/patients. That is the first fear (HCP). - Health professionals subjected to additional COVID-19 related tasks, patient flow decreased due to emerging concerns and fears of contracting the disease (HCP). - I have postponed my follow up at that time for fear of acquiring the disease from health professionals/health centers. The same is true for other clients in our area and some mothers have received their visit in private clinics as we perceived almost all staff were infected (W). - Health workers wear face masks for themselves, but they don’t let all clients wear face masks during facility visits (W). |
| Service deprioritized | - As much as possible we tried to make faster service provision for their children and give advice for them not come back frequently, they can manage themselves at home if it is easy (HCP) - We also used tele medicine for mild cases, because at the initial phase there was a direction of avoiding hospital visits for cases other than emergency (HCP). - Initially priority was given for patients who have cough but without compromising maternal and child health care services (HCP). |
| Low transportation access | - It is also another common reason for all of us to reduce client flow to the facility (W). - Initially …… mothers were staying at hospital unnecessarily due to absence of transportation/ambulance/ (HCP). - In this area there was no transport restriction, but numbers were reduced to half sit and cost was doubled. It was one of the factors to reduce flow (HCP). - Travel restrictions are also another reason for low client flow which is more pronounced among mothers from far kebeles (HCP). |
| Public panic | - At the beginning of covid-19 occurrence, the community panicked and feared acquiring the disease (HCP). - Our basic challenge is fear of the disease. The community heard the severity of the disease in the developed country in the media, but now the problem is solved (HCP). - Nationally the people panicked so there was a tendency of not visiting hospitals (HCP). - The community has been frightened of contracting the disease at the beginning (W). |
